# Did institutional racism contribute to adverse COVID-19 clinical outcomes in ethnic minority healthcare staff? Systematic review

**DOI:** 10.1101/2024.06.11.24308764

**Authors:** Oluwatomisin Adewole, Kismet Lalli, Holly Peters, Chris Price, Blanche Lumb, Micaela Gal, Alison Cooper

## Abstract

**Introduction:** Of the United Kingdom National Health Service (NHS) healthcare workers who died during the first wave of the pandemic, 63% belonged to an ethnic minority background, despite making up 21% of the NHS workforce. Previous research has considered biological and social causes such as obesity or overcrowded housing. This review aims to explore whether elements of institutional racism contributed to ethnic disparities in adverse COVID-19 clinical outcomes among healthcare staff.

**Method:** Eleven databases were searched including MEDLINE and the Cochrane Library. Eight healthcare organisations within the grey literature were also searched. A narrative synthesis was conducted.

**Results:** 20 studies were included for review. There were ethnic disparities in the rate of COVID-19 infection, mortality and wellbeing. Three elements of institutional racism were identified associated with these adverse outcomes, namely, overrepresentation of ethnic minority staff in frontline roles, discriminatory redeployment and harassment and bullying.

**Conclusion:** The pandemic exacerbated pre-existing racial inequalities within the UK healthcare workforce. Further research is required to clarify the definition of institutional racism and increase understanding of how this manifests in a healthcare setting and can be mitigated.

**Funding statement:** The Wales COVID-19 Evidence Centre was funded for this work by Health and Care Research Wales on behalf of Welsh Government.

## Introduction

Healthcare workers in the UK were seven times more likely to be exposed to the Sars-Cov-2 virus and develop COVID-19 compared with the general population during the COVID-19 pandemic (1). However, staff who identify as ethnic minority faced even greater adverse clinical outcomes due to events occurring in the first-wave of the pandemic (2). The first 11 National Health Service (NHS) doctors to die due to contracting the virus were of an ethnic minority background (3). By April 2020, 95% of all NHS doctors who died were minority ethnic (3). Furthermore, 63% of all NHS healthcare workers who died were Black, Asian or minority ethnic despite making up just 21% of the NHS workforce (3). Ethnic minority individuals were also more likely to be admitted into Intensive Care Units (ICU) and require invasive ventilation compared to their white counterparts (4).

Proposed biological causes of these disparities include higher genetic susceptibility to the coronavirus and the presence of pre-existing conditions (5). Other research places greater focus on socio-economic factors such as low-income employment, residing in over-crowded areas where the virus is more prevalent and transmissible, as well as culturally-influenced health-seeking behaviours (6). When these social and biological factors are accounted for in research, a disparity in outcomes still remains between ethnic groups, suggesting that there is another cause that is not being addressed (7).

One cause to consider is that of institutional racism. The British Medical Journal (BMJ) defines institutional racism as ‘the processes of racism that are embedded in laws (local, state, and federal), policies, and practices of society and its institutions that provide advantages to ethnic groups deemed as superior, while differentially oppressing, disadvantaging, or otherwise neglecting ethnic groups viewed as inferior’(2). Elements that comprise this term are outlined in Figure 1. This term is not clearly defined in research and other used definitions such as Macpherson’s(8), are less applicable to a healthcare context.

**Figure 1-.**
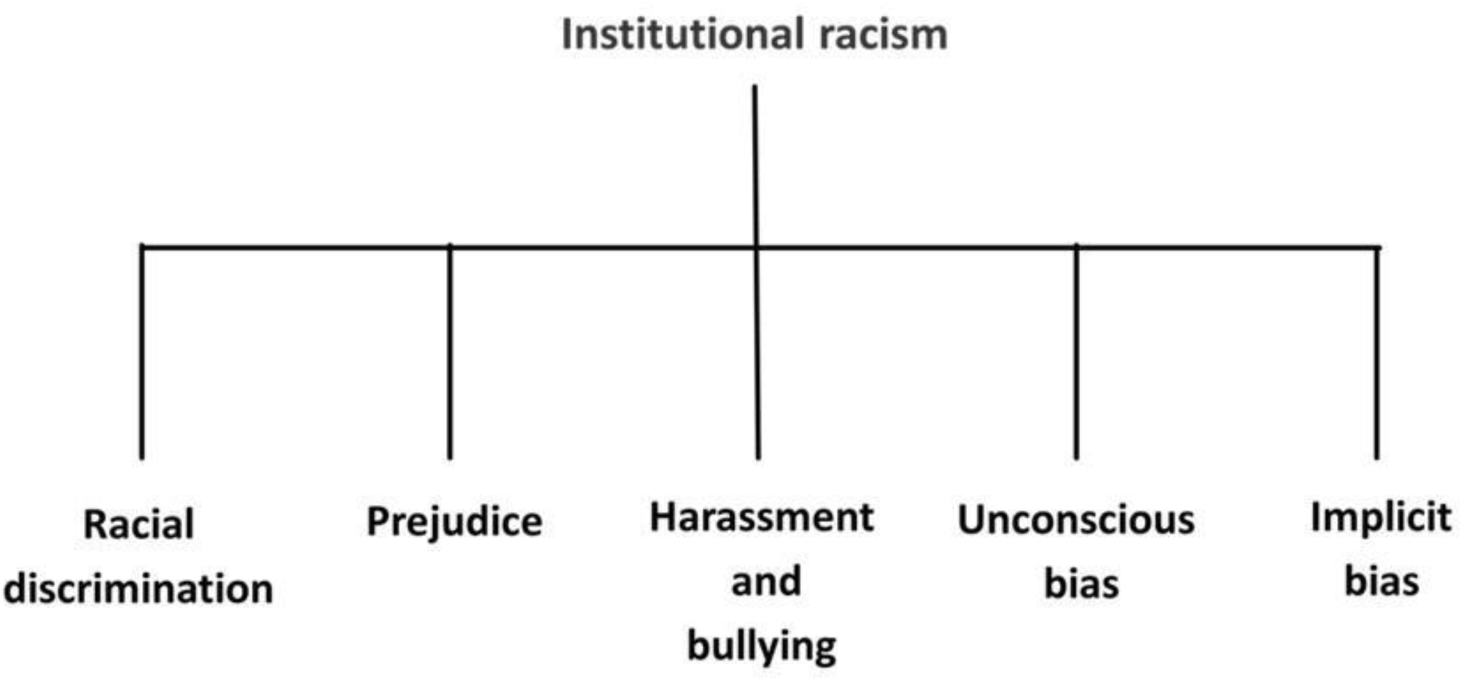
Elements of institutional racism (adapted by author) 159×68mm (150 x 150 DPI)

The aim was to explore whether racism persisting at an institutional level within the UK healthcare system contributed to the racial disparities in COVID-19 adverse clinical outcomes among NHS healthcare staff.

Our objectives included:

Identifying and describing the adverse clinical outcomes experienced by ethnic minority healthcare staff compared with white healthcare staff during the COVID-19 pandemic; Identifying factors that contributed to the differences in adverse clinical outcomes; Exploring how elements of institutional racism might contribute to these differences

## Method

We conducted this review in September 2021 accordance with the Cochrane Handbook of Reviews and the Preferred Reporting Items for Systematic Reviews and Meta-Analyses (PRISMA) guidelines (9). We registered the protocol with International Prospective Register of Systematic Reviews (PROSPERO)(10). We were supported by representatives from Health Education Improvement Wales (HEIW) and the General Medical Council (GMC) as stakeholders who had an interest in this area. They supported the review throughout: clarifying terminology, protocol planning, review progress meetings and dissemination of findings.

### Search strategy

We developed a search strategy, developed by stakeholders (CP, BL, KL, RJ) and two information scientists (EG, DM). Key search terms were divided into five concepts based on the research question: “COVID clinical outcomes”, “ethnic minority”, “healthcare staff/employees” and “institutional racism”.

### Sources of data

We searched eleven databases: MEDLINE, EMBASE, Cochrane Library, Web of Science, PsychINFO, SCOPUS and Joanna Briggs Institute Evidence-Based Practice database (JBI EBP), L*OVECovid, VA- ESP*, Cochrane COVID review bank and Collabovid. The searches occurred between 11_th_ – 30_th_ January 2022.

We also searched the grey literature between 30_th_ January – 5_th_ February 2022. Eight healthcare organisations were searched for grey literature material (Figure 2).

**Figure 2-.**
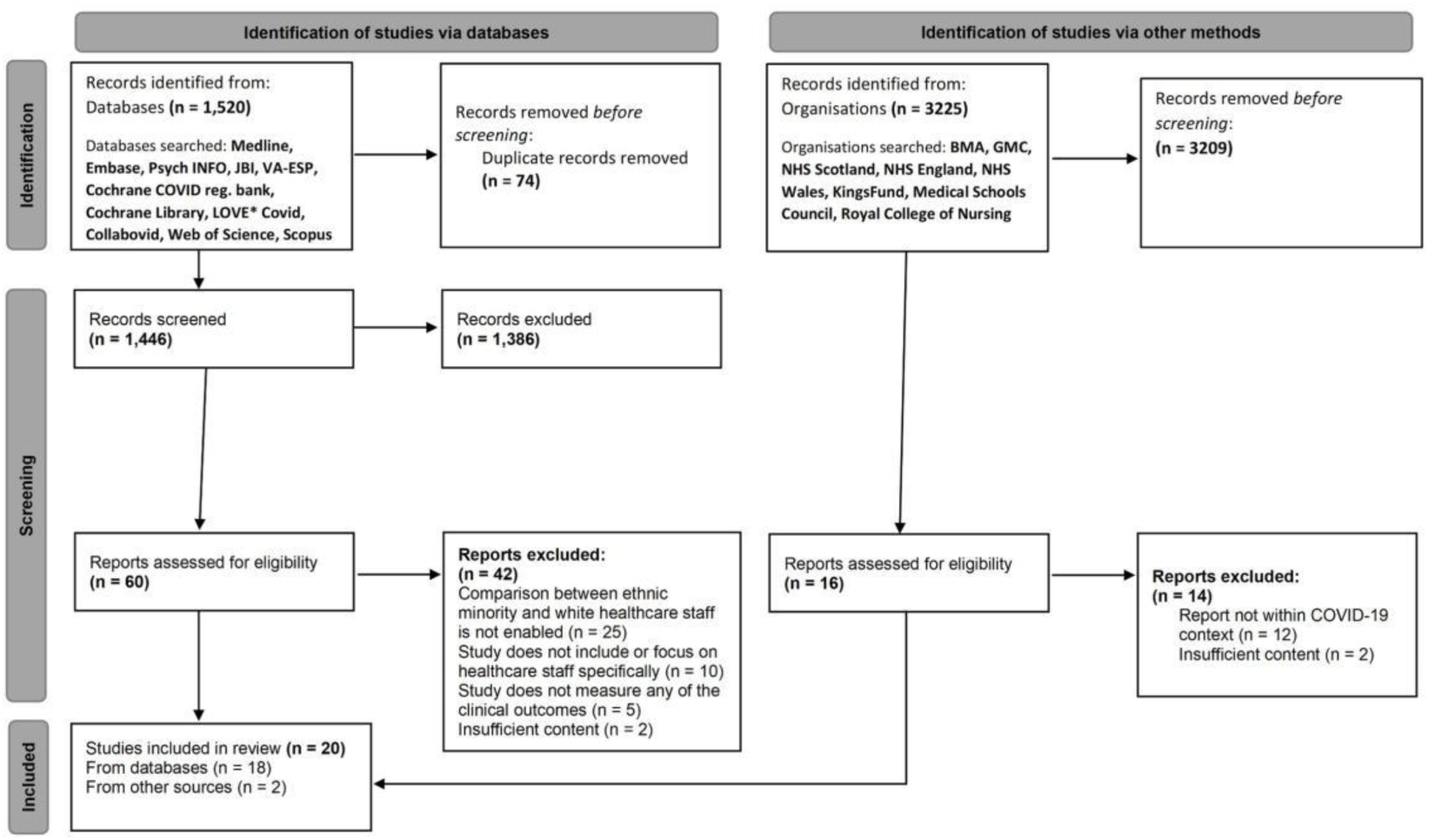
PRISMA Flowchart 259×149mm (220 x 220 DPI)

The eligibility criteria are outlined in Table 1. Clinical outcomes outlined in Table 1 were derived from current research in the general population and are not specific to healthcare staff and employees. Therefore, studies that assessed other adverse outcomes, for example wellbeing, were included.

**Table 1-.** Eligibility Criteria for inclusion and exclusion of studies 159×81mm (220 x 220 DPI)

|  | Inclusion | Exclusion |
| --- | --- | --- |
| Population | Healthcare staff/employees in the UK | Studies that do not include staff or only focus on patients and the general population<br>Non-UK based |
| Exposure | **Primary:** Institutional racism<br>**Secondary:** Socioeconomic background, pre-existing conditions, age etc. | Studies that do not report ethnicity |
| Comparison | Comparison of COVID-related adverse clinical outcomes between ethnic minority and white healthcare staff | No comparison enabled (based on ethnicity) |
| Outcome(s) | Rate/Incidence/Prevalence of Sars-Cov-2 or Covid-19 infection<br>Mortality<br>ICU admission<br>Hospitalisation | Studies that do not assess for these outcomes in ethnic minority staff |

### Study Selection

The studies identified, were independently screened by a single reviewer (OA) against the eligibility criteria. Studies were screened by title and abstract, followed by full text screening. 20% of the studies were double coded by a second reviewer (KL) to check the consistency in interpretation of the eligibility criteria. Any uncertainty was discussed among the research team.

### Data Extraction

Data were extracted from each eligible study by an independent reviewer (OA) using a data extraction form and presented in a Microsoft Excel spreadsheet. The data extraction table of all eligible studies is provided in the supplementary material. Data were extracted based on their sample characteristics, study design and the outcome measured (11).

### Critical Appraisal

We critically appraised each study according to their design (12). Four studies were appraised using the Joanna Briggs Institute (JBI) opinion and commentary checklists (13). Four studies used the JBI prevalence checklists (14). One study was appraised using the Critical Appraisal Skills Programme (CASP) qualitative checklist(15). Six studies were appraised using the Specialist Unit for Review Evidence (SURE) cross-sectional checklist and five were appraised using the SURE cohort study checklist(16, 17).

## Results

A total of 1520 studies was identified from electronic databases, and 1446 studies were screened by title and abstract following deduplication (Figure 2). 1386 studies were excluded by the reviewer (OA) as they were non-UK based, irrelevant to the PECO and beyond the context of the COVID-19 pandemic.

3225 items were identified from grey literature, press articles and blogs were excluded, 16 reports were assessed against the eligibility criteria and two reports were included. 20 studies remained for analysis, 18 sourced from electronic databases and two from the grey literature.

Most of the studies were observational in design (n = 11). This included six cross-sectional and five cohort studies. Four of the included papers were opinion pieces, whilst four others were surveys. One study was qualitative. Eighteen studies measured a COVID-19 clinical outcome among healthcare staff. Six of these also assessed for an element of institutional racism. Two opinion pieces discussed elements of institutional racism but no clinical outcome.

The included studies were mapped according to adverse COVID-19 clinical outcomes and the elements of institutional racism (Figure 3). This map, which is initialled according to study design, highlights many evidence gaps. Several studies assessed for more than one element of institutional racism or clinical outcome.

**Figure 3-.**
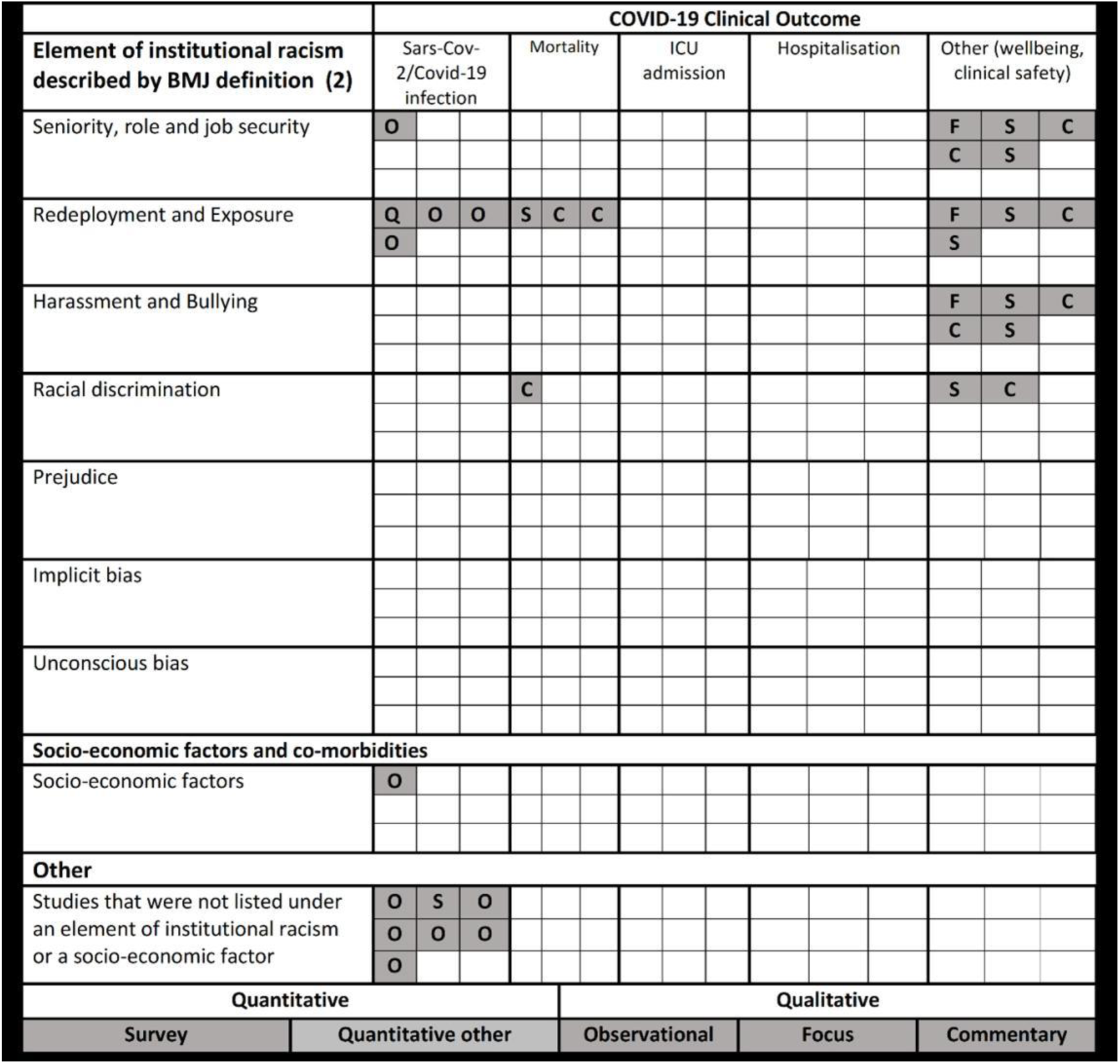
Evidence map of the included studies, mapped according to study design 398×378mm (87 x 87 DPI)

### Strength of Evidence

Most studies were of moderate-to-low quality due to poor justification of their methods and discussion of results. Many of the studies did not adjust for confounders when calculating the risk of COVID-19 infection, nor did they address the likelihood of information bias among participants.

There was a consistent lack of explanation regarding sample size or recruitment methods in the observational studies.

Four studies were of high quality; two cross-sectional, one cohort and one qualitative study (18–21). These studies obtained large datasets, provided thorough explanations of methodology and acknowledged sources of bias. The qualitative study provided a variety of rich, in-depth experiences of ethnic minority staff during the pandemic and a thorough thematic analysis. Critical appraisal of each study is provided in the supplementary material.

#### Ethnic disparities in adverse COVID-19 clinical outcomes

When socio-economic and demographic factors were adjusted for, an ethnic disparity among staff still remained (20, 23, 25). Other factors that may have contributed to ethnic disparities included genetic predisposition and inhabiting socially deprived regions (20, 24, 25).

##### SARS-CoV-2/COVID-19 infection

Of 10 studies that measured the prevalence of SARS-CoV-2 infection among ethnically diverse healthcare staff, nine found disparities in the prevalence, rate and risk between ethnic groups (18, 20–28). Eight studies, which measured the SARS-CoV-2 seroprevalence (detection of the SARS-CoV-2 antibody which indicates a positive infection) within the healthcare population, found that this was greater among ethnic minority than white healthcare workers (18, 20–24, 26, 27). One paper, which measured the seropositivity (presence of a positive SARS-CoV-2 antibody test) among healthcare staff, found no difference between ethnicities (28).

##### Hospitalisation and ICU admission

This outcome was measured in one paper, which found no difference in hospitalisation and Intensive Care Unit (ICU) admission rates between ethnic groups of healthcare workers (22).

##### Mortality

One cohort study examined the risk of COVID-related death among an ethnically diverse cohort of consultants, which found that the risk of death was greater among ethnic minority groups (29). When comparing white physicians with Asian, Black and Mixed physicians, the risk of mortality for ethnic minority women over 60 and men over 50, was two-fold. This level of risk increased five-fold in ethnic minority men over 60 (Hazard Ratio: 5.46, 5.50, 5.65 vs HR: 3.82) (29).

##### Mental and physical wellbeing

Six articles identified ethnic disparities in wellbeing, including clinical safety during the pandemic (19, 30–34). Ethnic minority staff were more likely to receive inadequate PPE-training whilst working on COVID wards and have reduced access to PPE equipment (30, 31). Ethnic minority nurses had concerns about inadequately designed masks, which they felt did not consider cultural or physiological differences between ethnic groups (33).

#### Potential elements of institutional racism

None of the studies used the term ‘institutional racism’. Discrimination and racism were alluded to by authors but un-defined (25, 27, 28). The elements of institutional racism identified in this review were derived from the BMJ definition(2).

##### Overrepresentation in frontline roles and lack of job security

> *‘Minority ethnic groups are systemically over-represented at lower level of NHS grade hierarchy, working in the shadow of snowy white peaks’(35)*

There is evidence of ethnic disparities in healthcare professions across six papers. Ethnic minority staff are more likely to work in junior roles, occupy lower pay bands and are underrepresented in senior, managerial clinical and non-clinical positions (20, 31–33, 35, 36). Two papers suggested that the pandemic exacerbated concerns over career progression among ethnic minority healthcare staff and increased the lack in promotion opportunities (19, 32). In a survey, members of staff reported feeling as though they were excluded from senior-level discussions concerning their safety whilst working during the pandemic and that they were appointed ‘riskier responsibilities’ in comparison with their white colleagues such as having to see patients without PPE (31).

Ethnic minority healthcare staff are more likely to undergo frequent employment changes due to overrepresentation in temporary roles (31, 35, 36). Ethnic minority staff were less likely to receive in-person training than their white colleagues whilst working in COVID wards (31).

Ethnic minority staff, particularly within the nursing profession, expressed feeling a lack of job security (19, 31, 33). An article reported that ethnic minority nurses felt as though they had a lack of agency or power when speaking to their line managers and senior staff (33).

##### Discriminatory redeployment and higher risk of exposure

> *‘I was told to see a COVID patient without extra PPE…when I expressed my concerns, I was dismissed and told I had no right to refuse to see any patient…I felt so undervalued and worthless’*(*37*)

Redeployment is the process of reassigning staff and employees to a new role or work location to meet new demands (38). Five studies found that ethnic minority healthcare workers were more likely to be redeployed to COVID wards than their white colleagues and were at greater risk of exposure to the virus (30, 31, 33–35). Both ethnic minority senior and junior healthcare workers were four times more likely to be redeployed to patient facing roles than their white colleagues (31). Ethnic minority staff believed themselves to be at greater risk of mortality because of ‘racially motivated’ redeployment methods (30, 33).

##### Harassment and bullying

> *‘There were…people who…chose to not follow guidance and they wanted to see patients face to face…because they were worried about their position in the team and about bullying’*(*19*)

A finding across five studies was an inability for ethnic minority staff to raise concerns regarding their clinical safety due to fear of how it may impact their job (19, 30, 31, 33, 34). Some felt that there was a ‘culture of appeasement’ that was prohibiting raising concerns of welfare (31). Staff described a ‘disregard’ and lack of understanding regarding the experiences of ethnic minority employees in the workplace, causing a barrier in conversation with their white managers (19). In a survey around wellbeing of healthcare staff during the pandemic, 32% of ethnic minority healthcare workers felt overlooked and were treated without dignity or respect. 39% of respondents felt unsupported by their white seniors (34). They spoke of the ‘pressure’ applied to those working from home “because [they] were not trusted to be doing what [they] should be doing” by white managers (19). Ethnic minority staff were reluctant to work from home due to fear of perceived special treatment and how this might impact interprofessional relationships at work (19).

## Discussion

### Main findings

Among 20 UK-based studies, ethnic disparities were found in the acquisition of COVID-19 infection, clinical wellbeing and mortality rates. Potential evidence of institutional racism included: overrepresentation of ethnic minority staff in frontline roles, discriminatory redeployment, and harassment and bullying, which may have contributed to these ethnic disparities in outcomes.

### Strengths and Limitations

This is the first systematic review to assess the impact of institutional racism impacting UK ethnic minority healthcare staff during the pandemic. We searched 11 databases and extensively in the grey literature. Selecting exclusively UK-based studies increases the transferability of the findings to the NHS. Stakeholder involvement helped to focus the review and interpret findings.

Most evidence was of moderate-to-low quality. The data extraction and critical appraisal processes were not checked by a second reviewer due to time constraints, this could have introduced information bias. Additionally, interpretations of elements of institutional racism may have been subjective. However, this was mitigated through discussion of uncertainties within the research team and stakeholders. Another limitation was a lack of consistency regarding ethnic categories, where attention to specific sub-groups varied between studies.

### Comparison with literature

Before the pandemic, ethnic minorities were disproportionately represented in high-risk, low-paying jobs within the NHS (39–41). This was exacerbated during the pandemic, which saw an uptake of at-risk healthcare roles by predominantly ethnic minority staff (42). These roles are less-valued within the healthcare system and have been associated with job insecurity, poorer health and a greater risk of exposure (42, 43).

Wider evidence suggests that this overrepresentation may be due to underemployment of ethnic minority staff in senior positions (44). White staff are more likely to be appointed into senior roles from shortlisting compared with ethnic minority staff (45). Additionally, ethnic minority staff constitute 10% of NHS trust boards despite making up over 19.7% of the workforce (45, 46). This excludes them from employment decisions, hinders career progression and provides unequal opportunities to staff.

A consistent finding identified within this review was a lack of agency and job insecurity felt among NHS staff. Literature attributes this to multiple factors, including the feeling of inferiority to white colleagues, which is perpetuated by the presence of pay-gaps and ongoing workplace racism. As a result, staff members have reported feeling unable to raise concerns with their seniors regarding their wellbeing, for fear of how this will impact their job. Ethnic minority staff are 1.5 times more likely than their white colleagues to enter the formal disciplinary process due to referral by a senior member of staff (47, 48). This creates a workplace culture of fear and appeasement, in which staff feel compelled to dismiss safety concerns to avoid being perceived as a ‘troublemaker’ (49). Pope et al. referred to this culture as a form of ‘organizational silence’ designed to prohibit ethnic minority staff from voicing out (49).

This review highlighted the exacerbation of harassment and bullying towards ethnic minority healthcare staff during the pandemic. The results align with a pre-pandemic picture of harassment and bullying in the NHS. Staff from ethnic minority groups have consistently been subjected to higher levels of bullying, harassment and abuse from their colleagues, compared with white staff(45). Despite the actions of organisations such as the Workforce Race Equality Standard (WRES), harassment and bullying towards ethnic minority staff is still increasing. In 2021, the NHS staff survey revealed that the rate of bullying and harassment of ethnic minority staff had increased from 14.5% in 2019 to 17% (50). However, as the NHS is an employer organisation, their surveys may be subject to reporting bias, as ethnic minority staff may feel uncomfortable disclosing their personal experiences (51). Furthermore, bullying and harassment appears to be most prevalent in nursing, where 1 in 5 nurses are of an ethnic minority background (52). This may be due to a greater exposure to vertical harassment and bullying, from staff in senior positions, as well as horizontal harassment and bullying, from staff within the same level (53). The consequences of harassment and bullying include a deterioration in mental wellbeing and poorer health outcomes as a result of chronic stress (51, 54, 55). It is apparent that the fear of being stereotyped or disrespected by their colleagues deprives ethnic minority staff of an environment where they feel able to voice concerns about their safety.

Prejudice was not explicitly stated in any of the studies, nor has it been sufficiently defined in wider literature. However, we identified an association between discrimination and the review findings. Several of the studies allude to the redeployment process as a form of discrimination against ethnic minority healthcare staff (31, 33, 35, 37). The BMJ states that discrimination results in ‘inequitable access to opportunities and resources’(2), this review identified two studies in which there was a clear disparity in PPE and training opportunities between white and ethnic minority healthcare staff (31, 37). Similarly, the lack of ethnic minority representation in senior NHS positions presents as ‘disadvantaging or otherwise neglecting’ (2) these groups as they are excluded from decisions involving their employment and safety (19, 31, 32).

## Conclusion

The pandemic highlighted pre-existing racial inequalities present within the UK healthcare workforce. These inequalities need to be addressed by governing bodies and healthcare organisations, with interventions robustly evaluated. Further research is required to clarify the definition of Institutional racism and increase understanding about how this manifests in a healthcare setting and can be mitigated.

## Data Availability

All data produced in the present study are available upon reasonable request to the authors

## Acknowledgements

The authors of this review would like to thank Professor Adrian Edwards, Rhian Jones and Katie Laugharne for their support and feedback.

## Ethical Approval

None Required

## Search plan strategy

**Research project title:** Did institutional racism contribute to adverse COVID-19 clinical outcomes in ethnic minority healthcare staff and employees? A Systematic Review

**Databases searched:** MEDLINE, EMBASE, Cochrane Library, Web of Science, PsychINFO, SCOPUS, JBI EBP, L*OVECovid, VA-ESP*, Cochrane COVID review bank and Collabovid

**Key concepts and search terms:**

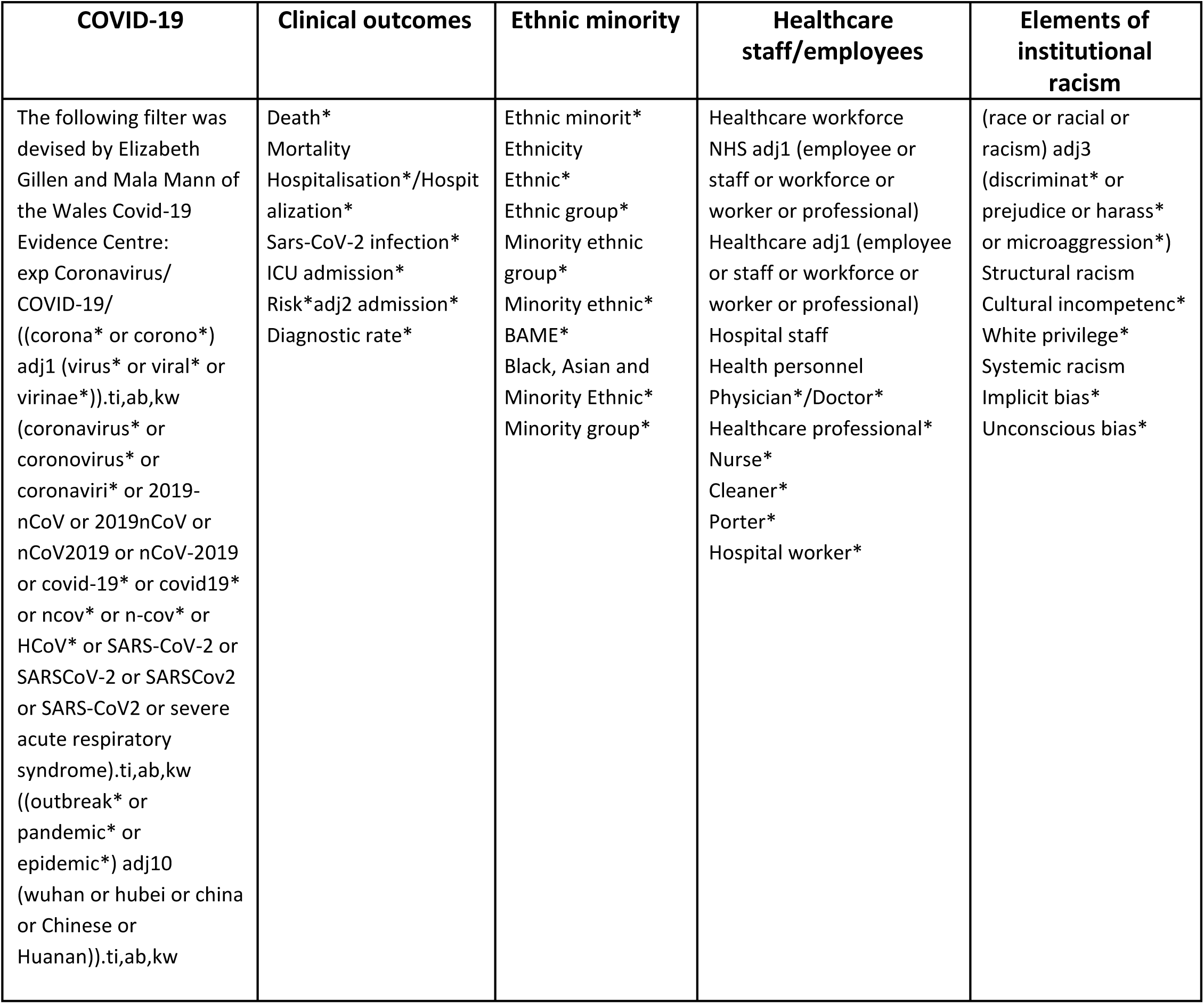

## Supplementary material: Data Extraction tables (**1–3**)

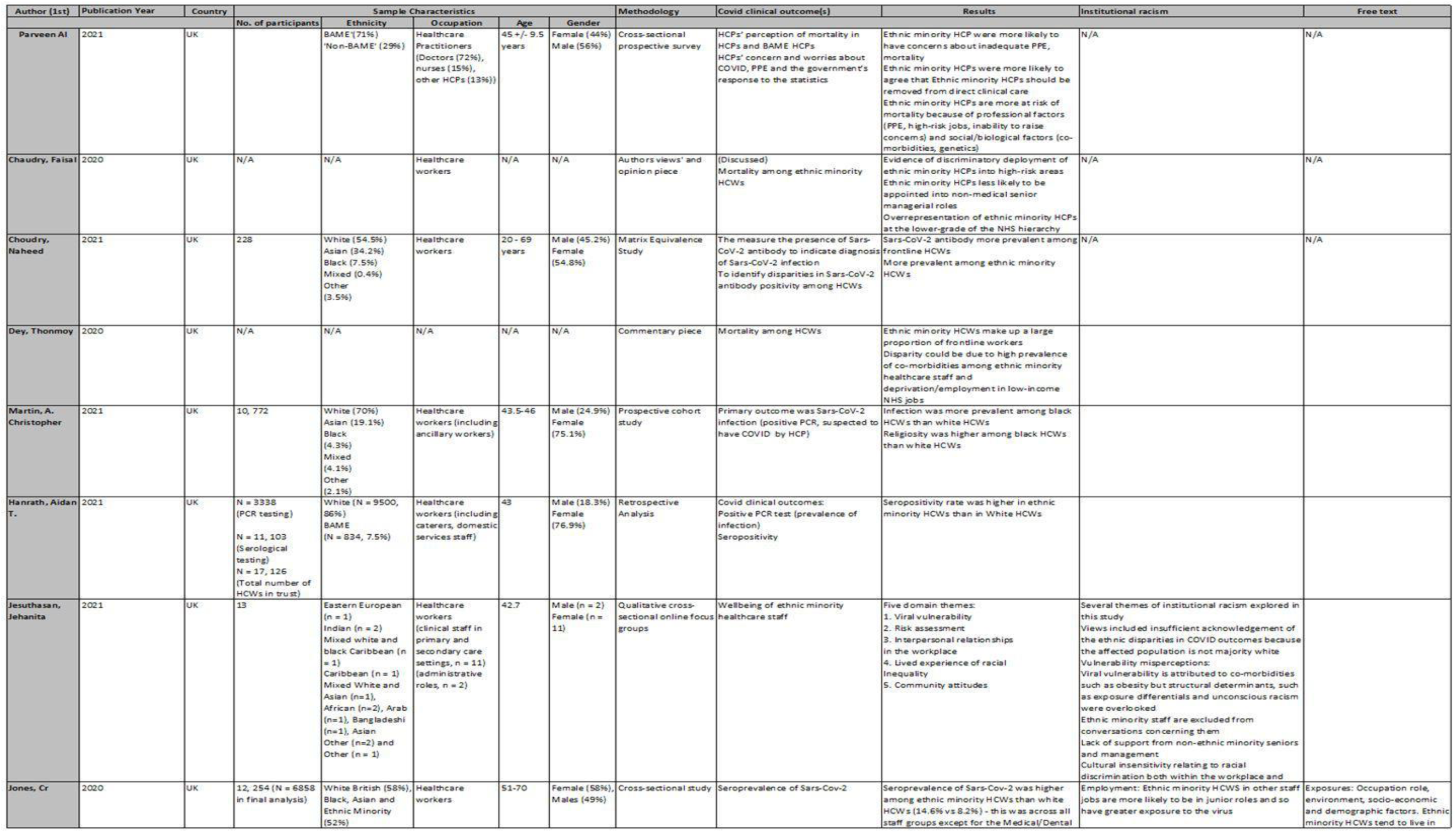
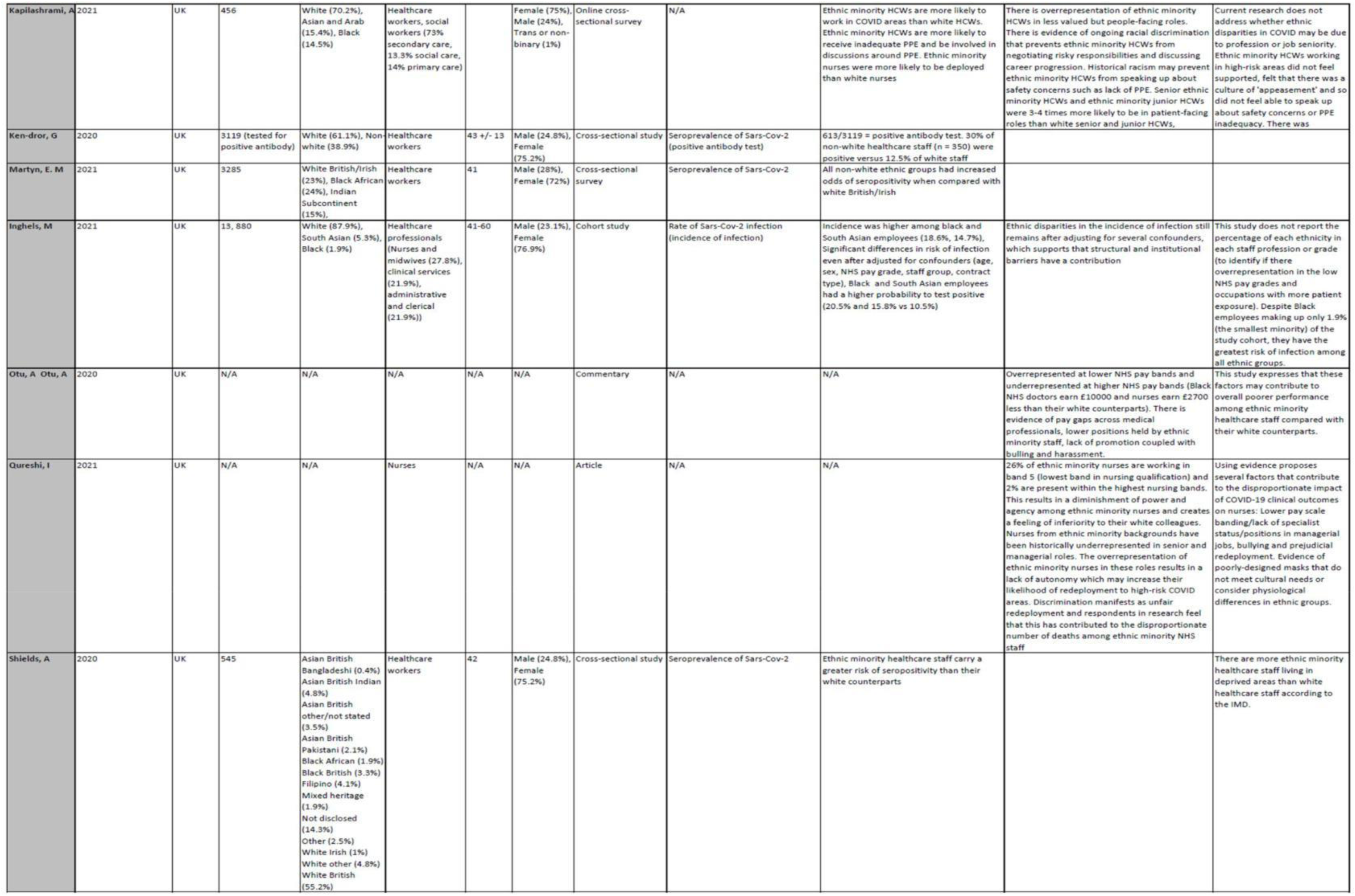
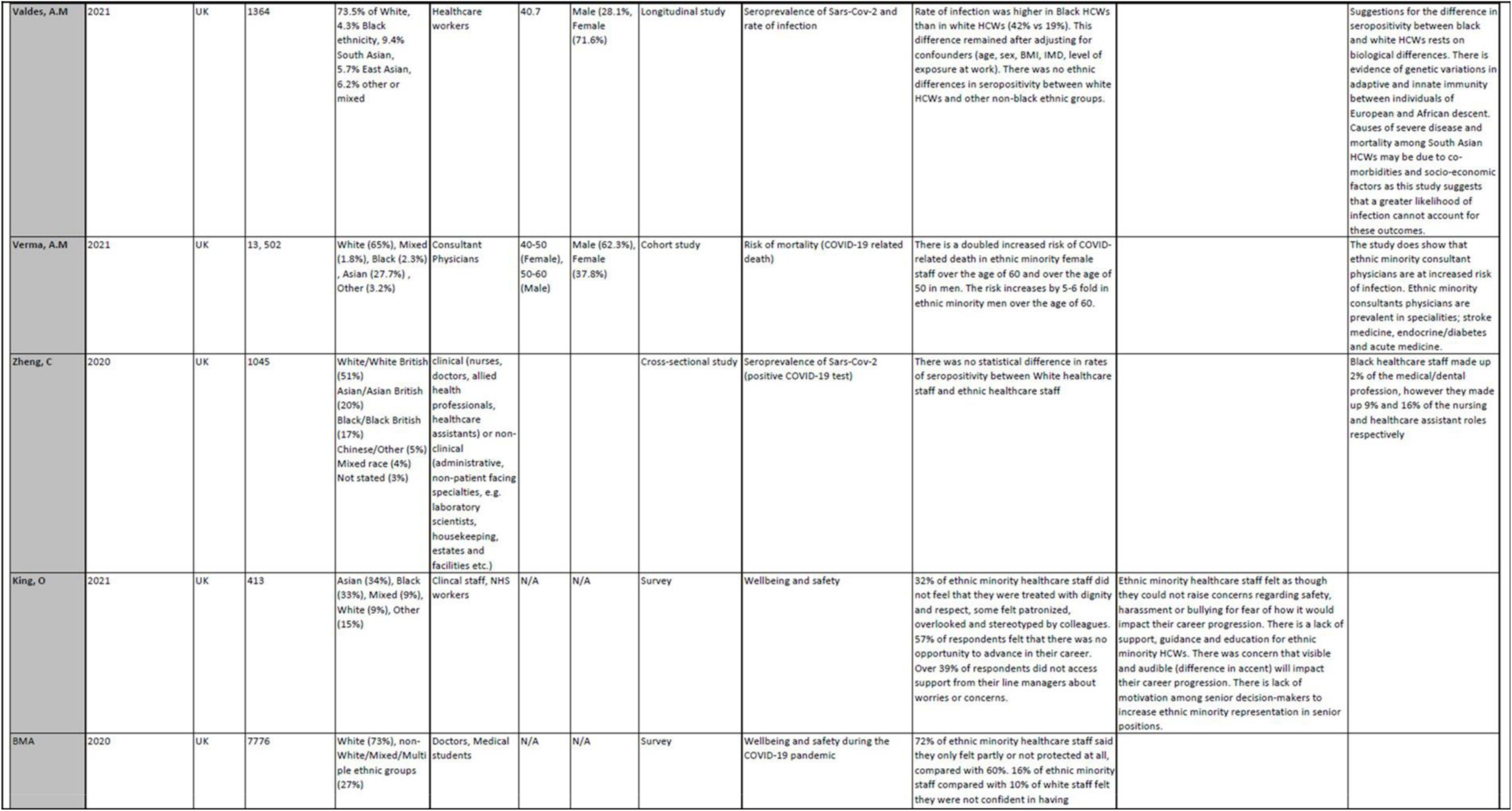

## Supplementary Material: Critical Appraisal Tables (**1–12**)

Supplementary Material: Critical Appraisal Tables (1-12)
| Specialist Unit for Review Evidence (SURE) cohort checklist |  |  |  |  |  |
| --- | --- | --- | --- | --- | --- |
| <u>Author,</u><br><u>Year, Country of Publication</u> | **Martin, A. Christopher**<br>**et al.**<br>**2020, United Kingdom** | **Inghels, M et al.**<br>**2021, United Kingdom** | **Verma, A.M et al.**<br>**2021, United Kingdom** | **Valdes, A.M et al.**<br>**2021, United Kingdom** | **Choudhry, N et al.**<br>**2021, United Kingdom** |
| **Is the study design clearly stated?** | Yes – The authors state in the introduction and method section that this is a prospective nationwide cohort study | Yes – It is stated both in the title and the methodology that this study is a occupational cohort study | No – The study design is not clearly stated in either the introduction or methods section however this was deciphered from other details of the methodology | Yes – It is stated to be a longitudinal study | Can't tell – The description given in the study design is one of a cohort study, however it is not clearly stated by authors what exactly the study design is. There are two study's occurring however it is does not meet the criteria of a mixed-methods approach. |
| **Does the study address a clearly focused question?** | Yes – The study aimed to address how demographic and occupational factors influence infection risk in healthcare staff | Yes – The aim of this study was to quantify the prevalence of infection among ethnic minority healthcare staff and to decipher a causal pathway by considering other factors | Yes – The authors aim to determine those at increased risk of dying for COVID among a cohort of consultant physicians | Yes – Aims to address ethnic differences in risk of Sars-Cov-2 infection among healthcare workers | Yes – Aims to address disparities in Sars-Cov-2 infection in healthcare workers |
| **Are the setting, locations and relevant dates provided?** | Yes - The study is set in the UK context and is nationwide, so there is no specific location, however the authors explain this.<br>Recruitment was via a questionnaire that was administered between December 2020 and March 2021 | Yes – The study is set at three hospital trusts within Lincolnshire. Participants were followed retrospectively between 1<sup>st</sup> January 2020 – 10<sup>th</sup> February 2020 | Yes – The study is set nationwide and is looking at data collected from consultants in the England, Scotland and Wales through Royal College of Physicians (RCP). The data being analysed was collected from a questionnaire distributed on the 30 September 2018 (the last available census). | Yes – The study is set across 5 hospitals in the UK (Nottingham and London); Nottingham City Hospital and Queen's Medical Centre - both Nottingham University Hospital NHS trust; COVID-sortium, London: St Bartholomew's, Nightingale and Royal Free Hospitals<br>The study took place between the 23<sup>rd</sup> March – 10<sup>th</sup> July 2020 | Yes – The study is set in Barts Health trust in East London. The recruitment took place between June – August 2020 |
| **Were participants fairly selected?** | Yes – The authors list the eligibility criteria in the Methods section. Participants over the age of 16, employed by any one of the UK Regulatory bodies (i.e. General Medical Council) and working as a healthcare worker (HCW) or ancillary worker (cleaner, porter etc.) were eligible to participate in this study | Yes – The authors clearly outlined the eligibility criteria in the methods section of this study. All employees of the three trusts were selected to follow (excluding students as they do not have a staff assignment group). Employees working in multiple facilities were also excluded. | Can't tell | Partial Yes – Healthcare workers including nurses, doctors, allied professionals and frontline workers were recruited from both Nottingham and London. However, other workers such as ancillary and admin staff were only recruited from London | Yes – Eligibility criteria is provided by the authors in supplementary material. |
| **Are participant characteristics provided?** | Yes – A summary table of the characteristics of each cohort (total cohort and analysed cohort) are provided which list demographic, household and occupational factors | Yes – A summary table is provided listing demographic and occupational characteristics | Yes – A summary table is provided listing the age, sex and ethnicity of the cohort | Yes – Provided in a table alongside their serology status | Yes – A summary table is provided including demographic details and work status |
| **Are the measures of exposures & outcomes appropriate?** | Yes<br>Outcome: **Sars-Cov-2 serology** which was assessed by either self-reported positive PCR or PCR testing performed by the research team<br>Exposure: Ethnicity which was self-reported by participants which was categorised into five broader groups | Yes<br>Outcome: Positive PCR test result (collected from human resources data within each trust) | Can't tell | Yes<br>Outcome (seropositivity): PCR and antibody testing<br>Exposures (demographic and occupational factors): Data collection from the respective trusts/hospitals and self-reported by participants | Yes<br>Outcome (Sars-Cov-2 antibody): Matrix Equivalence study (a quantitative measure of calculating seropositivity), antibody testing |
| **Was bias considered? e.g. recall or selection bias** | Yes – Authors state that there is a potential for selection bias given the amount of information that is self-reported, i.e. the self-reported PCR test result, demographic and occupational information. They state that self-reporting of outcomes has the potential to cause recall bias, however they do not expect this to have a substantial effect on their outcome measure | Can't tell – The authors state that the quality of data collection was overseen by senior healthcare staff at every stage of the study, however there is no clear statement that considers any form of bias | No | Yes – There were no differences between the timing of infection among seropositive (infected) HCW, which the authors state may be due to recall bias. They also consider a risk of symptom recall bias among HCWs. They were able to adjust for confounders which may have reduced bias. | No – Authors did not clearly outline whether they had considered bias |
| Is there a description of how the study size was arrived at? | No | No – Although it is implied that this size is based off of the number of employees within each of the trusts | No | No – Though it is implied that the size is based off of the number of employees who had been infected with the virus / had symptoms | Yes – A sample size and power calculation was included |
| Are the statistical methods well described? | Yes – These methods are well described and clear. | Yes – They clearly state the regression analysis that they used in adjusting for confounders and other variables | Yes | Yes | Yes |
| Is information provided on participant flow? | Yes – Figure 1 outlines the formation of the analysed cohort included at which point and the number participants were excluded (including reasons for exclusion) | No | No | No | Yes – outlines each stage of the study, recruitment and number lost to follow-up (including the reasons) |
| Are the results well described? | Yes – Findings are presented using confidence intervals and p-values. Categorical variables are summarised as percentage and frequency. Comparison between ethnic group of demographic and occupational factors was done using chi-square tests and Kruskal-Wallis tests. | Yes – Confidence intervals used to present data but not consistently | No – Results presented using only hazard ratios and percentages. No presentation with confidence intervals. | Yes – Results presented using 95% Confidence intervals and p-values, as well as odd ratios. | Yes – Data presented with confidence intervals, odds ratios and percentages |
| **Is any sponsorship/conflict of interest reported?** | Yes – Some of the authors have substantial roles in organisations that may have vested interest in the subject matter of this study. One of the authors is a director of the University of Leicester Centre for Black Minority Ethnic Health and another is the medical director for the GMC | No | No | Two of the authors declared their involvement with external organisations such as Medical Research Council, COVID- 19 Rapid Response Rolling Call but other authors had nothing to declare | The study received funding from Abbott Rapid Diagnostics |
| **Did the authors identify any limitations and, if so, are they captured above?** | Yes – Limitations were mainly recorded around selection and recall bias due to self-selection and reporting by participants. They are captured above. | Yes – Authors identify variables and confounders that they were not able to adjust for in their analysis and also discuss the low generalisability of the study | Yes – Underestimation of risk due to not accounting for confounders such as co-morbidities | Yes – Clearly stated limitations due to recall bias, sensitivity/specificity of testing and difficulties in adjusting for confounders | Yes – Did not adjust for certain confounders (socio-economic inequalities) which may have linked ethnicity with sars-Cov-2 serology status |
| **Overall quality grade** | High | Moderate | Low | Moderate | Moderate |

| Specialist Unit for Review Evidence (SURE) cross-sectional checklist |  |  |  |  |  |  |
| --- | --- | --- | --- | --- | --- | --- |
| Author, Date and Country of Publication | Parveen Al, et al, 2021, United Kingdom | Hanrath, Aidan T, et al, 2021, United Kingdom | Jones, Cr, et al, 2020, United Kingdom | Ken-Dror, Gie et al., 2020, United Kingdom | Shields, Adrian et al., 2020, United Kingdom | Zheng, C et al, 2020, United Kingdom |
| Is the study design clearly stated? | Yes – The study design is a cross-sectional prospective study that has collected data through an online questionnaire | No – The authors have cited the study design to be a ‘Retrospective Analysis’ however there are no details provided of what this entails. From the introduction the reviewer can decipher that this a retrospective cross-sectional study design | Yes – This is clearly stated in the methodology | Yes – It is stated in the abstract and methods section that it is a cross-sectional study | Yes – Authors state that this is a cross-sectional study in the ‘Design’ section | No – The design of this study is not explicitly stated anywhere in the text |
| Does the study address a clearly focused question? | Yes – This study aims to describe the perceptions of risk and mortality associated with COVID-19 among ethnically diverse healthcare staff | Yes – The study explores the prevalence of PCR positivity among healthcare staff and the potential association between seropositivity and demographic/occupational factors | Yes – To explore the relationship between socio-demographic factors (i.e. ethnicity) and differences in seroprevalence among healthcare staff | Yes – This study sought to quantify the prevalence of Sars-Cov-2 in healthcare workers | Yes – This study aims to measure how many participants are infected with Sars-Cov-2 or have positive Sars-Cov-2 serology | Yes – The reviewer is able to decipher the PECO from the introduction, however it is not clearly outlined |
| Are the setting, locations and relevant dates provided? | Yes – Information relevant to the study was provided. The questionnaire was conducted online via Google Forms between 28<sup>th</sup> April – 4<sup>th</sup> May 2020 | Yes – The study was set in Newcastle-Upon-Tyne (NUTH) hospitals trust. The dataset analysed was from between 10<sup>th</sup> March – 6<sup>th</sup> July 2020 | Yes – The study’s setting is North Bristol NHS trust. Data was collected from participants between January – June 2020 and antibody testing took place in May 2020 | Yes – This study is set in Surrey in two separate hospitals (Ashford and St Peters) | Yes – This study was set in Birmingham at the UHBFT (University Hospitals Birmingham NHS Foundation trust) and recruitment took place over a period of 24 hours | No – The testing period is vaguely outlined with no specific dates mentioned. Authors state vaguely that the study is set in a teaching hospital in London (a huge city, in which there are |
|  |  |  |  |  | between 24<sup>th</sup> – 25<sup>th</sup><br>April 2020 | multiple teaching hospitals) with no information regarding the demographics or population it serves etc. (if there is a reason for this omission of detail, it is not given) |
| **Were participants fairly selected?** | Can't tell – Participants had to be a healthcare professional working in the UK. Participants were contacted through various social media sites and healthcare professional networking sites (i.e. Doctors. Net, National Primary Care) which may have excluded those who do not use social media. 71% of the sample are described as 'BAME', which may indicate selection bias. | Yes – All National Health Service (NHS) employees in the NUTH trust were selected as participants, which may minimise selection bias. The eligibility criteria is not stated clearly in this study, however this may be due to the fact that the only essential criteria is to be an active employee of this particular NHS trust. | Yes – All employees of the NHS trust were invited to participate in the study. Though eligibility criteria is not clearly outlined and the participation in antibody testing was voluntary. | All adults employed by the trust were invited for antibody testing, there is no clearly outlined eligibility criteria except for participants needing to be >18y/o. | Can't tell – Participants were individually selected, they were invited on the basis that they were employed by the trust. Participants were excluded if they showed COVID symptoms on the day or if they had been self-isolating for 2 weeks prior | Can't tell – No information given on how participants were enrolled (was it self-reported, where they invited etc.)<br>No eligibility criteria clearly outlined<br>Staff who were symptomatic were referred by their line managers to drive-through testing pods which could introduce bias |
| **Are participant characteristics provided?** | Yes – A table is provided that details the ages, genders and professional groups of the sample. Ethnicity is | Yes – Two tables summarising occupational and demographic characteristics are provided in supplementary material | Details regarding the characteristics of the participants are provided in results (to explain findings), | Yes – A summary table of key sample characteristics is provided | Yes – A summary table of sample characteristics is provided (Age, Sex and Ethnicity) | No – There is a table provided that details the ethnicity of the participants but no other demographic |
|  | recorded as either 'BAME' or 'non-BAME' |  | however there is no summary table provided that clearly outlines the characteristics of the sample cohort |  |  | characteristics which is not sufficient |
| **Are the measures of exposures & outcomes appropriate?** | Can't tell – Questionnaire is used to collect data on perceptions around COVID-19 related risk and mortality. The survey included a combination of Likert scales (1 – 10) and multiple choice questions | Yes – Outcomes: Seropositivity – PCR/antibody testing, data Hospitalisation/ICU admission/ventilation – Dataset Exposures (occupational factors, i.e. role and demographic factors, i.e. deprivation) are measured using data from ESR (Electronic staff record) and postcodes | Yes – Outcomes: Antibody testing and employee medical records Exposures (characteristics): Age, sex, employee status - Employee medical records Deprivation - IMD | Yes Outcomes: Sars-Cov-2 prevalence – Antibody testing results Exposures (Characteristics): - Self-reported medical records | Yes Outcomes: Sars-Cov-2 serology – antibody testing and nasal swab Exposures: Self-reported data and records from NHS England | Yes Outcomes: Sars-Cov-2 serology – PCR testing and occupational health data/staff records Details on measures of exposure are not clear |
| **Is there a description of how the study size was arrived at?** | No | Yes – The study size was decided based on the number of staff employed by the NHS trust at the time of analysis which was 17, 126 | Yes – The size was based on the number of staff employed by the North Bristol NHS trust | No – However it is implied that the study size is the number of individuals who responded to the invite for testing (4000 were invited for antibody testing, 3119 responded) | No – Authors stated that there was no predefined sample size as participants self-reported for enrolment | No |
| **Are the statistical methods well described?** | No –The use of SPSS was described and comparison performed using t-test. Data presented as 95% CI and | Yes | Yes Data was presented using descriptive analysis with n (%) or median (IQR) | Yes – They are described in depth including how they will be summarised and presented (i.e. | Yes | No – Statistical methods are not described |
|  | ORs, but no justification is given |  |  | descriptive analyses of data is summarised with standard deviation or median (IQR)) |  |  |
| **Is information provided on participant eligibility?** | Yes – Any individual who works as a healthcare professional in the UK is eligible | Yes – There was no exclusion criteria, the main inclusion criteria was that the participant was employed by the NUTH NHS trust. HCWs not directly employed by the trust and whose information was not available in ESR were excluded. | No – There was no outlined or specific eligibility criteria provided for the reviewer's discretion, however the authors do state that every employee of the trust is eligible including temporary and bank staff | Can't tell – Eligibility criteria is not specific or clearly outlined for the reviewer, however in the text the authors do state that all staff over the age of 18 were invited for testing | Yes – The authors briefly state criteria for exclusion from the study (mentioned previously) | No |
| **Are the results well described?** | Yes – The confidence intervals and standard deviation are provided, along with percentages and summarised tables of figures | Yes – The authors describe the results of each outcome assessed and the association (if any) between the outcome and the demographic/occupational factors 95% Confidence intervals and odds ratios are provided to present the data | Yes – Any differences in seroprevalence between staff groups based on key characteristics (ethnicity, deprivation score) are outlined | Yes | Yes – They are described in detail using statistical analyses (95% Confidence intervals and adjusted Odds ratios), however it could be more clearly or logistically laid out | No - There is vague evidence of thorough statistical analyses and no use of confidence intervals or p-values to summarise data |
| **Is any sponsorship/conflict of interest reported?** | No | No | No | No | No | No |
| **Did the authors identify any limitations and, if so, are they captured above?** | No – They briefly mention that there is selection bias but not in sufficient detail. They vaguely mention that they have similar limitations to other rapid online surveys but do not go on to list these, which underlines the transparency of the results | Yes – The authors did identify limitations within their methodology and study design as well as how they presented their results | Yes – The authors identify that voluntary participation in antibody testing may have introduced selection bias into their study. | Yes – Confounders such as underlying health conditions, BMI or deprivation indices were not adjusted for<br>This study only used antibody testing to measure seroprevalence which is not as thorough without PCR testing alongside to determine seropositivity of Sars-CoV-2 | Yes – Introduction of selection bias due to self-enrolment of participants. | Yes – Lack of available data and vague eligibility criteria regarding COVID symptoms etc. |
| **Overall quality grade** | Moderate | High | High | **Moderate** | **Moderate** | **Low** |

| Critical Appraisal Skills Programme (CASP) qualitative checklist |  |
| --- | --- |
| <u>Author,</u><br><u>Year, Country of Publication</u> | Jesuthasan, Jehanita et al.,<br>2021, United Kingdom |
| **Was there a clear statement of the aims of the research?** | Yes – To gain insight into the impact of COVID-19 on the mental wellbeing of ethnic minority healthcare staff **the** |
| **Is a qualitative methodology appropriate?** | Yes – It is conducive to exploring in-depth the thoughts, experiences and opinions of the participants in a way that quantitative methodology could not achieve |
| **Was the research design appropriate to address the aims of the research?** | Can't tell – This study utilised semi-structured focus groups however the justification for this design is not explicitly stated. The authors do mention that focus groups will enable participants to respond to each other's opinions and comments which might be helpful in exploration of their overall experiences. It was a cross-sectional design which is appropriate given the context of the study, it enables the researchers to get a snapshot sample of the experiences of ethnic minority healthcare and social workers during the pandemic |
| **Was the recruitment strategy appropriate to the aims of the research?** | Yes – The authors state that they use purposeful sampling to recruit participants and this was done to select participants who were likely to meet the objectives of the study (i.e. from an ethnic minority background individuals, working in a healthcare/social setting). This recruitment was done through public and patient involvement networks as well as through advertisement in NHS trusts. |
| **Was the data collected in a way that addressed the research issue?** | Yes – Data collection was via focus groups. The authors did not clearly state why this method was chosen, however they did provide details regarding the form of data |
| **Has the relationship between researcher and participants been adequately considered?** | Yes – The facilitator of the both focus group sessions was non-ethnic minority, which they argued may have led to some bias/inhibited conversation, however they countered this by explaining that the facilitator had substantial experience in diverse fields and low-middle income countries. Additionally, another author, who is an ethnic minority, was present in both sessions |
| **Have ethical issues been taken into consideration?** | Yes – Details regarding the purpose and method of the focus group were reiterated to the participants. They were assured that they were welcome to withdraw from the focus group at any time and were encouraged to ask questions at any stage. |
| **Was the data analysis sufficiently rigorous?** | Yes – Thematic analysis was used as analysis. The themes identified are outlined clearly in a diagram. Each theme is explored and explained in considerable depth and detail for the reviewer. |
| **Is there a clear statement of findings?** | Yes – There are several statements included in the discussion that clearly summarise the evidence found |
| **How valuable is the research?** | The authors state that these findings are supported by other literature and studies into the mental wellbeing of ethnic minority healthcare staff. They highlight gaps in consideration of ethnic disparities in COVID outcomes, i.e. structural barriers, unconscious bias and express the need for more supportive management |
| **Overall quality grade** | High |

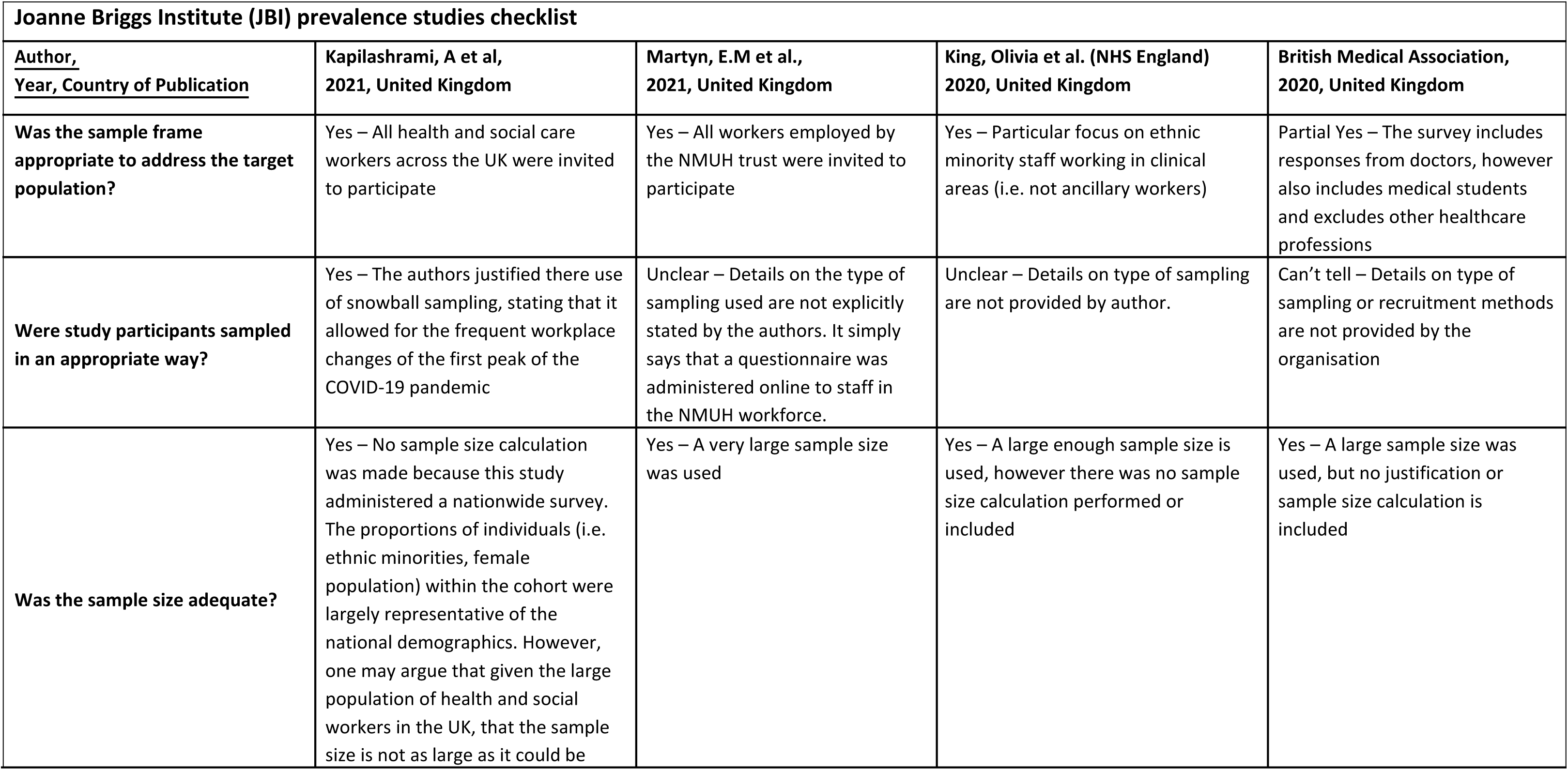
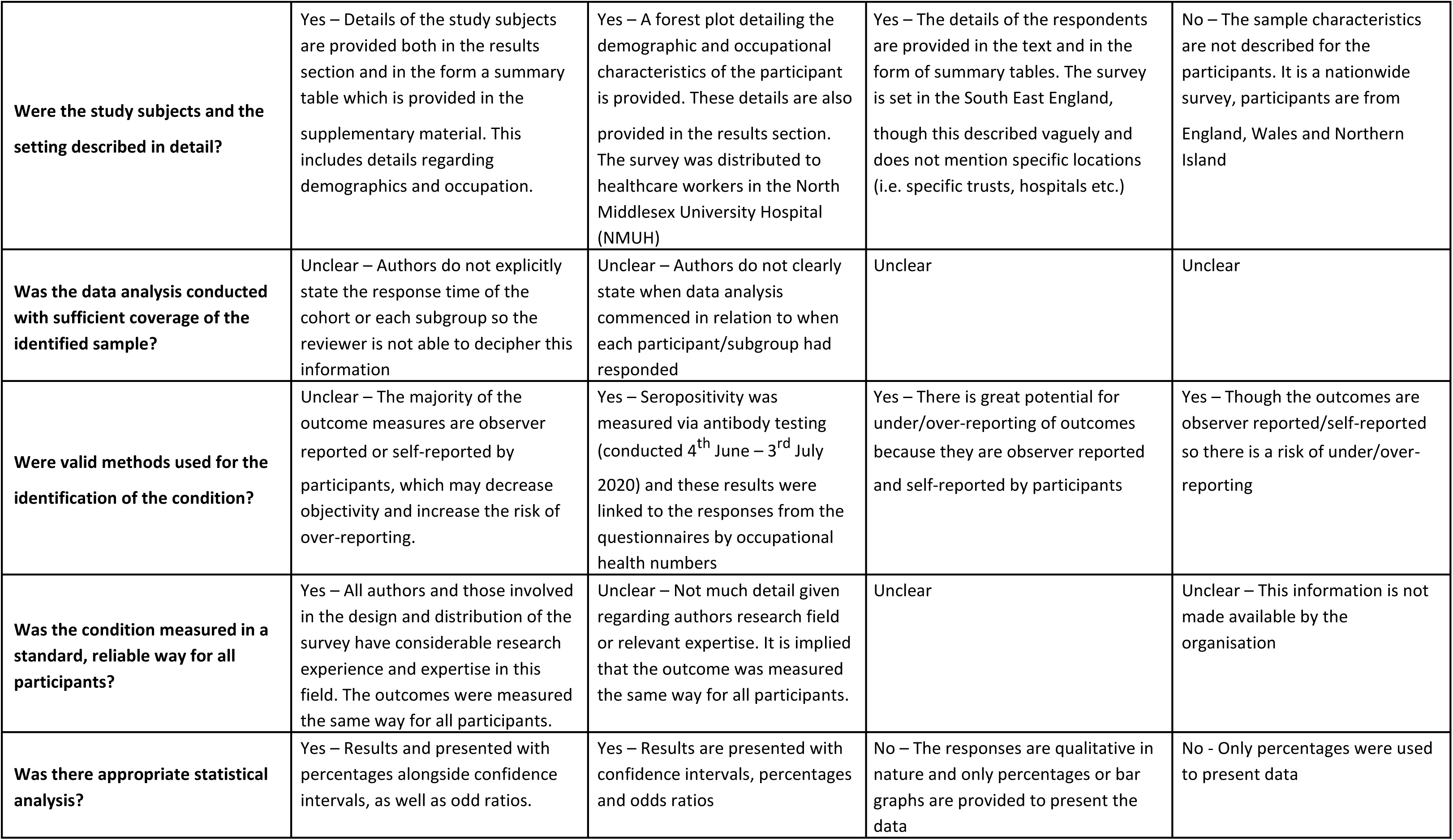

| Joanna Briggs Institute (JBI) opinion and commentary checklists |  |  |  |  |
| --- | --- | --- | --- | --- |
| Author, Date, Country of Publication | Chaudry, Faisal et al.<br>2021, United Kingdom | Dey, Thonmoy et al<br>2020, United Kingdom | Otu, Akaninyene<br>2020, United Kingdom | Qureshi, Irtiza<br>2021, United Kingdom |
| Is the source of the opinion clearly identified? | Yes – The author is named and credited | Yes | Yes | Yes |
| Does the source of opinion have standing in the field of expertise? | Yes – The publications and credits of the author are stated within the text and there is evidence of expertise in global health | No – No publications or credits are cited for any of the authors of this commentary which undermines the validity and expertise of the source | Yes – The publications of each of the authors are accessible and there is evidence of expertise in infectious diseases | Yes – Publications and credentials are accessible for the reviewer |
| Was the response rate adequate, and if not, was the low response rate managed appropriately? | Yes – Response rate was good and the majority of people who received the survey responded. | Unclear - The authors do not comment on the response rate, but the majority of staff invited to complete the survey, did respond which implies an adequate response rate | Unclear – The authors only state the number of respondents but do not state the number of participants that were invited to complete the survey | Unclear – No information regarding the total number of participants invited to complete the survey |
| Overall quality grade | Moderate – High | Moderate – Low | Low | Low |
| **Are the interests of the relevant population the central focus of the opinion?** | Yes - The subject of this text is 'COVID-19 and BAME healthcare staff' and the content revolve around the disproportionate impact of COVID health outcomes on 'BAME' healthcare staff and discusses the potential causes (overrepresentation of ethnic minority HCWs in front line roles) | Unclear – The title of this commentary implies that the focus is on the relationship between ethnicity and COVID-19 with no specificity to healthcare workers. However, much of the content of the text focuses on the factors underlying high mortality and cases among ethnic minority healthcare workers rather the general population. A clearer statement would be appropriate. | No – Ethnic minority HCWS are not the central focus (the general ethnic minority population is discussed in this commentary), however they are discussed in depth in part of the text | Yes – This commentary focuses specifically on ethnic minority nurses and the disproportionate effect of COVID clinical outcomes on this population |
| **Is the stated position the result of an analytical process, and is there logic in the opinion expressed** | Yes – The authors studied data relating to the COVID-19 mortality among healthcare workers and analysed this against population demographics | No – No analytical process has been described or can be implied from the text however, the arguments expressed are logical and presented in a critical way | No - However there is logic in the expressed opinion | Yes – The argument presented is very logical and based on wider literature |
| **Is there reference to the extant literature?** | Yes – However the inclusion of external literature is non-critical and exists only to support the argument being made. It is not non-biased nor critically appraised. Having said this they do discuss residing in urban areas as a confounding factor. | Yes – External literature is referenced in a descriptive way, however it is non-critical and does not strengthen the argument being made. | Yes - External literature is referenced to support the presence of systemic discrimination that silences ethnic minority HCWs | Yes – The majority of this opinion piece is supported by wider literature which |
| **Is any incongruence with the literature/sources logically defended?** | No – The authors draw on external literature to show that there is wider support for their main line of argument but no inconsistencies are identified | No | No | No |
| **Quality grade** | Moderate – High | Low – Moderate | Moderate | Moderate – High |

